# Validation of *International Classification of Diseases, 10^th^ Edition, Clinical Modification (ICD-10-CM)* Code I68.0 for Cerebral Amyloid Angiopathy

**DOI:** 10.1101/2024.01.16.24301394

**Authors:** Samuel S. Bruce, Santosh Murthy, Hooman Kamel

## Abstract

**Introduction:** Cerebral amyloid angiopathy (CAA) is a common cause of intracerebral hemorrhage and cognitive impairment in the elderly. Though definitive diagnosis requires post-mortem pathological analysis, clinical and radiographic criteria allow for noninvasive, in vivo diagnosis. We sought to validate the new ICD-10-CM diagnostic code for CAA with respect to the recently updated Boston criteria, version 2.0.

**Methods:** We conducted a retrospective study of inpatient and outpatient encounters at a single hospital center, Weill Cornell Medicine (WCM), from 10/1/2015 to 12/31/2018. We randomly selected 25 encounters with the ICD-10-CM code I68.0 and 25 encounters with a primary diagnosis of intracerebral hemorrhage or ischemic stroke, without code I68.0. A single board-certified neurologist, blinded to ICD codes, reviewed detailed medical records and images from brain MRI and CT scans from all 50 selected encounters and identified subjects with possible or probable CAA by Boston criteria 2.0. Sensitivity and specificity of ICD-10-CM code I68.0 was calculated.

**Results:** Of the 50 selected encounters, 21 (42%) met criteria for possible or probable CAA: 18 (36%) met criteria for probable CAA, 2 (4%) met criteria for probable CAA with supporting pathology, and 1 (2%) met criteria for possible CAA. The ICD-10-CM code I68.0 was found to have a sensitivity of 81% (95% CI, 58-95%) and specificity of 72% (95% CI, 53-87%) for identifying possible or probable CAA.

**Conclusion:** The ICD-10-CM code I68.0 was found to have good sensitivity and moderate specificity for CAA as defined by current clinical and radiographic diagnostic criteria. Based on our results, this code may be useful for identifying patients with CAA in future research using administrative claims data.

## Introduction

Cerebral amyloid angiopathy (CAA) is a small vessel vasculopathy characterized by accumulation of amyloid beta in vessel walls.^1^ It is common in the elderly population, with prevalence increasing with age,^2^ and is an independent risk factor for non-traumatic intracerebral hemorrhage (ICH) and cognitive impairment.^3-5^ Though CAA can only be definitively diagnosed with tissue analysis from autopsy, clinical and radiographic criteria have been formulated to allow for noninvasive in-vivo diagnosis.^6^ The most recent update of these criteria, Boston criteria version 2.0, introduced non-hemorrhagic magnetic resonance (MRI) markers to existing hemorrhagic markers and clinical criteria, increasing sensitivity with regards to the gold standard of histopathological diagnosis.^6,7^Until recently, research using administrative claims data in CAA has been limited due to the lack of diagnostic codes for the disease. A new diagnostic code for CAA, I68.0, was introduced with the *International Classification of Diseases and Related Health Problems, 10*^*th*^ *Edition-Clinical Modifications (ICD-10-CM)* in 2015. However, to our knowledge, this code has not been validated with respect to existing clinical and radiographic criteria. Thus, the objective of our study was to assess the validity of *ICD-10-CM* code I68.0 in identifying patients who meet Boston criteria 2.0 for CAA.

## Methods

We conducted a retrospective cross-sectional study of inpatient and outpatient encounters at a single hospital center, Weill Cornell Medicine (WCM), a tertiary referral center in New York City, from 10/1/2015 (the date ICD-10 was introduced in the US) to 12/31/2018 (the latest date of our ICH and ischemic stroke registry). We randomly selected 25 encounters with the *ICD-10-CM* code I68.0 for CAA and 25 encounters for ICH or ischemic stroke. A single board-certified neurologist (S.S.B.) reviewed detailed medical records and images from MRI and computed tomography (CT) scans from all 50 selected encounters. Clinical features of CAA (spontaneous ICH, transient focal neurological episodes, and cognitive impairment or dementia) were determined by detailed chart review. Assessment of MRI markers of CAA (lobar ICH, lobar microhemorrhage, convexity subarachnoid hemorrhage [SAH], cortical superficial siderosis [CSS], multispot white matter hyperintensities [WMH], and centrum semiovale dilated perivascular spaces [PVS]) was performed as previously described.^8^ Determination of possible or probable CAA was based on Boston criteria version 2.0.^6^We used standard descriptive statistics including percentages for dichotomous variables and medians with interquartile ranges (IQR) to describe demographic and clinical variables. Using the Boston criteria 2.0 as the gold standard, we calculated the sensitivity and specificity and 95% confidence intervals (CI) of *ICD-10-CM* code I68.0. This study was approved by the WCM Institutional Review Board, which granted a waiver of informed consent.

## Results

In the study cohort, 25 patients (50%) were female, and the median age was 75.5 (IQR 68-80.75). Blinded adjudication identified 21 encounters (42%) as meeting criteria for possible or probable CAA: 20 (40%) met criteria for probable CAA, with 2 of the 20 meeting criteria for probable CAA with supporting pathology and 1 (2%) meeting criteria for possible CAA. Clinically, 8 (38%) subjects with possible or probable CAA presented with cognitive impairment or dementia, 6 (29%) presented with ICH, 5 (24%) presented with transient focal neurological episodes, and 2 (10%) presented with convexity SAH. Upon detailed chart review, 4 (19%) subjects with possible or probable CAA had a prior history of ischemic or hemorrhagic stroke.

Among 20 subjects with probable CAA, 5 (25%) had MRI evidence of lobar ICH, 10 (50%) had CSS or convexity SAH, 8 (40%) had multispot WMH, 5 (25%) had dilated centrum semiovale PVS, and 20 (100%) had at least one lobar microhemorrhage; the median number of microhemorrhages was 6.5 (IQR 3-49). The one subject that met criteria for possible CAA had a single small lobar ICH, with no other lesions. The ICD-10-CM code I68.0 had a sensitivity of 81% (95% CI, 58-95%) and specificity of 72% (95% CI, 53-87%) for identifying cases meeting Boston criteria 2.0 for possible or probable CAA.

## Discussion

In this retrospective cohort study, ICD-10-CM code I68.0 was found to have good sensitivity and moderate specificity in identifying patients meeting Boston criteria 2.0 for possible or probable CAA.

This study has multiple strengths. First, adjudication of cases was performed by a trained expert blinded to diagnosis code. Second, we included encounters in both the inpatient and outpatient setting, with a control group of patients with ICH and ischemic stroke that are likely to have similar clinical and radiographic features to CAA patients. Third, we used objective and well-validated diagnostic criteria for our gold standard.

Our study also has several limitations. Only one patient in the study fulfilled criteria for possible CAA, two patients included had supporting pathology, and no patients fulfilled criteria for definite CAA (which requires a full postmortem histopathological examination). Thus, the ability of code I68.0 to identify patients meeting these designations separately, without also including patients meeting criteria for probable CAA, is undetermined. Lastly, our study was only conducted in a single hospital center, and therefore, the generalizability of these results may be limited.

## Conclusion

We found that ICD-10-CM code I68.0 had a sensitivity of 81% and specificity of 72% for CAA as defined by current clinical and radiographic diagnostic criteria. Based on our results, this code may be useful for identifying patients with CAA in future research using administrative claims data.

## Data Availability

All data produced in the present work are contained in the manuscript.

## Disclosures

Dr. Bruce is supported by NIH/NCATS grant KL2-TR2385. Dr. Murthy is supported by the NIH/NINDS (K23NS105948).

Financial disclosures for Hooman Kamel: PI for the ARCADIA trial, which received in-kind study drug from the BMS-Pfizer Alliance for Eliquis and ancillary study support from Roche Diagnosics; Deputy Editor for *JAMA Neurology*; clinical trial steering/execuive commijees for Medtronic and Janssen; endpoint adjudicaion commijees for AstraZeneca, Novo Nordisk, and Boehringer Ingelheim; and household ownership interests in TETMedical, Spectrum Plasics Group, and Ascenial Technologies.

## Notes

### Author Declarations

Institutional Review Board of Weill Cornell Medicine gave ethical approval of this work, and granted a waiver of informed consent.

### Summary of Updates

We updated the results to include the presenting clinical features of patients with possible or probable CAA, and report the percentage of patients with possible or probable CAA that had a prior history of ischemic or hemorrhagic stroke. We also made a correction to the results - 5 patients (not 4) with probable CAA had MRI evidence of lobar ICH.

